# Protocol summary and statistical analysis plan for the randomized trial of early detection of clinically significant prostate cancer (ProScreen)

**DOI:** 10.1101/2023.05.09.23289669

**Authors:** Jaakko Nevalainen, Jani Raitanen, Kari Natunen, Tuomas Kilpeläinen, Antti Rannikko, Teuvo Tammela, Anssi Auvinen, the ProScreen Trial Team

## Abstract

**Introduction:** Evidence on the effectiveness of prostate cancer screening based on prostate-specific antigen is inconclusive and suggests a questionable balance between benefits and harms due to overdiagnosis. However, diagnostic accuracy studies have shown that detection of clinically insignificant prostate cancer can be reduced by magnetic resonance imaging combined with targeted biopsies.

The aim of the paper is to describe the analysis of the ProScreen randomized trial to assess the performance of the novel screening algorithm in terms of the primary outcome, prostate cancer mortality, and secondary outcomes as intermediate indicators of screening benefits and harms of screening.

**Methods:** The trial aims to recruit at least 111,000 men to achieve sufficient statistical power for the primary outcome. Men will be allocated in a 1:3 ratio to the screening and control arms. Interim analysis is planned at 10 years of follow-up, and the final analysis at 15 years. Difference between the trial arms in prostate cancer mortality will be assessed by Gray’s test using intention to screen analysis of randomized men. Secondary outcomes will be the incidence of prostate cancer by disease aggressiveness, progression to advanced prostate cancer, death due to any cause and cost-effectiveness of screening.

**Ethics and dissemination:** The trial protocol was reviewed by the ethical committee of the Helsinki University Hospital (HUS 2910/2017). Results will be disseminated in an international peer-reviewed journal(s) and at scientific meetings.

**Trial Registration:** NCT03423303

**STRENGTHS AND LIMITATIONS:**

- This population-based, randomized multicenter trial targeting at recruiting 111,000 men will provide high quality evidence on the effectiveness of a novel screening strategy for prostate cancer mortality
- Broad eligibility criteria and pragmatic approach embedded in normal clinical practice enhances the external validity of the trial and provide evidence applicable to decision making in public health and health care
- Challenges for the trial include the maintenance of high compliance to screening and the extent of opportunistic PSA testing in the population

## Introduction

Prostate cancer is the most common cancer in men in many industrialized countries and causes substantial mortality (Culp et al. 2020). Screening based on blood prostate-specific antigen (PSA) has been shown to decrease prostate cancer mortality, but the evidence from randomized trials is not conclusive (Hugosson et al. 2019, Pinsky et al. 2019). Systematic reviews of randomized controlled trials have concluded that PSA screening may at best lower prostate cancer mortality, but not all-cause mortality. However, the balance between benefits and harms was regarded as problematic due to frequent overdiagnosis, and complications from biopsies and overtreatment (Ilic et al. 2018, Fenton et al. 2018, Paschen et al. 2022).

Several studies have shown that detection of clinically insignificant prostate cancer can be reduced by magnetic resonance imaging (MRI) combined with targeted biopsies of the suspect foci, instead of systematic biopsies of the entire prostate (Schoots et al. 2015; Ahmed et al. 2017). However, previous studies have mostly focused on the diagnostic performance, i.e., cancer detection at a single evaluation. A hybrid screening/diagnostic study and a screening trial using MRI were recently published (Hugosson et al. 2022, Eklund et al. 2021).

Here we describe the analysis of the ProScreen randomized screening trial to assess the performance of a novel screening algorithm in terms of the primary outcome, prostate cancer mortality, and secondary outcomes, used as intermediate indicators of benefits and harms of screening. Following good statistical practice, this statistical analysis plan (version 1.0) was finalized prior to completion of recruitment and short-term follow-up data collection. It was written following the guidelines provided in Gamble *et al*. (2017) as applicable. Any unforeseen deviations from the plan will be described and justified carefully in the respective reports.

## Trial overview

### Trial design

The ProScreen trial is a population-based, randomized multicenter trial that investigates the effectiveness of a novel screening strategy combining PSA, a four-kallikrein panel, and MRI on prostate cancer (PCa) mortality over a 15-year period from randomization (Auvinen et al. 2017). The rationale is to minimize detection of clinically insignificant cancers, while maintaining a high sensitivity for aggressive cancers in order to reduce overdiagnosis without compromising mortality benefits. An interim analysis of PCa mortality is planned at 10 years of follow-up.

On 15 January 2018, the trial was registered at clinicaltrials.gov (NCT03423303). The ethical committee of Helsinki University Hospital reviewed the protocol (tracking no. 2910/2017). Permissions to collect data from health care registers was obtained from Finnish Institute for Health and Welfare (before the era of FinData, Dnro THL/676/5.05.00/2018). A written informed consent is provided by each participant in the screening arm.

Recruitment started in October 2018 and is still ongoing.

### Study population

All men aged 50–63 years (at the time of sampling of the trial population) with Finnish or Swedish as mother tongue residing in the trial municipalities constitute the trial population. Men with a prevalent prostate cancer will be identified through the Finnish Cancer Registry or hospital pathology databases and excluded.

We have identified for the trial the entire target population from the Digital and Population Data Services Agency, comprehensively without any sampling. The initial trial population consists of men residing in Helsinki and Tampere.

Currently, we are increasing the sample size by recruiting men also in the other municipalities within the Helsinki and Tampere metropolitan areas (Vantaa, Espoo, and Kauniainen, as well as Nokia, Lempäälä, Pirkkala, Ylöjärvi, and Kangasala with a total of 57,000 men in the target age group). The target population covers comprehensively all eligible men in the municipalities, both in the original Helsinki and Tampere areas and the new municipalities.

### Sample size

We estimated that we could find 110,000–120,000 men in the target age group based on the population projections from 2020 to 2034 from the ten municipalities (Statistics Finland 2021). We requested the overall number of deaths and the number of PCa deaths from Statistics Finland by age group from 1990 to 2019. The proportion of PCa deaths had barely changed at all during the 30-year period and hence, we based our sample size calculation on these figures. With a 1:3 random allocation to the screening arm relative to the control arm, we estimated that at least 240 PCa deaths would occur in the control arm during the first ten years of the trial, and at least 520 PCa deaths by 15 years of follow-up.

Assuming a relative hazard of 0.75 for the screening relative to the control arm, Schoenfeld’s formula indicates that an 80% power would be reached by a total of 506 PCa deaths (Schoenfeld 1983) with type I error rate set at 5%. Assuming a total of 650 PCa deaths – 520 in the control arm and 130 in the screening arm – the power of the study would be 89%. Hence, we aim at a final sample size of at least 111,000 men to ensure adequate statistical power and precision at the final analysis.

### Randomization and screening intervals

All eligible men will be randomly allocated to screening and control arms in a 1:3 ratio. Within the screening arm, re-screening interval is adapted by the baseline PSA:

- Men with initial PSA≥3 ng/ml are re-invited every two years,
- Men with PSA 1.5–2.99 ng/ml every four years, and
- Men with PSA<1.5 ng/ml after six years.

By the time of writing this plan, we have randomized 61,193 men with 15,299 allocated to the screening arm and 45,894 to the control arm. Analyses will compare the entire screening arm, regardless of the actual screening attendance and interval employed, to the control arm, unless otherwise specified.

Randomization list consists of batches of randomized men. The list is generated centrally by a designated study biostatistician at the coordinating unit, who maintains the documentation including program codes and the resulting lists include information of randomization dates, personal identification numbers (linkable to study ID number) and the arm allocated. Randomization lists are only shared confidentially to study personnel if needed for study conduct.

### Screening procedures

At every screening attendance, three consecutive tests are conducted in a stepwise manner before biopsy:

1. All participating men give a blood sample for determination of PSA at a local laboratory.
2. If the PSA is 3 ng/ml or higher, a four-kallikrein panel is analyzed from a second vial of plasma from the initial draw using an algorithm incorporating four proteins (total PSA, free PSA, intact PSA and human kallikrein-2) and age. The result is expressed as probability of a clinically significant PCa.
3. Men with both PSA≥3 and kallikrein score≥7.5% are referred to MRI. T2-weighted, diffusion-weighted and dynamic contrast-enhanced imaging is employed in accordance with the European Society for Urogenital Radiology guideline (de Rooij 2020). The findings are classified according to the Prostate Imaging Reporting and Data System (PI-RADS v2.1), which is a 5-point scale to combine the MRI findings and indicate the likelihood of a significant cancer. Scores of 3–5 indicate at least a suspect finding warranting directed biopsy.

Only targeted biopsies are employed, with 2–4 cores per region of interest depending on the size. Only screen-positive men with negative MRI but PSA density >0.15 undergo systematic biopsy as a safety measure (to avoid missing clinically significant cancers). Similar fusion-guided biopsy systems are used at the two trial sites and evaluated by experienced uropathologists using standardized procedures.

A random sample of screen-negative (on test steps 1 and 2) men are also invited to prostate MRI and asked to give blood, urine and stool samples in order to serve as a control group to estimate frequency of suspicious MRI findings in the general population, and as a reference group in analyses of biological samples.

### Protocol deviations

A tabular presentation of different types of protocol deviations along with their frequencies and percentages will be presented. Any protocol deviations detected after randomization will be carefully documented. Among them, men later found out not to have met the eligibility criteria at the date of randomization can be excluded from the analysis (post-randomization exclusions).

In the case of major protocol violations affecting a substantial proportion of men, separate per protocol analyses will be conducted to support the main analyses. In the screening arm, incomplete attendance, or compliance with the screening procedures is likely to occur. In the control arm, we will obtain data on contamination, i.e., mostly self-initiated PSA testing.

When considering unforeseen lack of compliance with the protocol, all means to ensure objectivity in the exclusion principles from per protocol analyses will be taken. Participants in both arms will be considered according to the same principles. Protocol deviations not related to the screening procedures are expected to appear in approximately 1:3 ratio for the arms. Obvious deviation from this ratio would be reported and interpreted as a potential source of bias.

### Blinding

Blinding in the conventional sense is not applicable: men are aware of being invited to screening. Hence, this is an open trial with screening and control arms.

Concrete measures to prevent bias, if any, from the awareness of the trial arm were nevertheless taken: (i) the control arm is blind to the fact that they are part of the trial; (ii) allocation concealment is ensured by the centralized randomization procedure preventing foreknowledge of upcoming arm allocation; and (iii) communication to the general public on trial is kept to the minimum to prevent contamination (e.g. by self-initiated PSA testing) among men in the control arm.

In addition, we underline that the primary outcome of the study, PCa death, is an objective outcome. The possibility of bias in its evaluation only relates to the assessment of the cause of death. The death certificates are filled by physicians with no involvement in the trial and can be assumed to be independent of trial arm, especially as deaths from prostate cancer are likely to occur years after the diagnosis and hence unaffected by detection through screening or other means. Importantly, a previous study within the ERSPC trial has shown that the cause-of-death data provided by Statistics Finland agreed almost perfectly with the assessment of a blinded expert panel in the Finnish center of the trial and was independent of the trial arm (Mäkinen et al. 2008, Kilpeläinen et al. 2016).

### Data collection process

Table 1 summarizes the stages of the data collection process, targeted participants, and information and samples obtained.

**Table 1.** Data collection process of the ProScreen trial.

| Process stage | Target population | Information collected | Samples collected |
| --- | --- | --- | --- |
| Baseline | Participants | Family history<br>Previous PSA and Bx<br>Generic QoL/utility (15D, EQ5D)<br>Out-of-pocket costs<br>PSA and four-kallikrein panel | Plasma<br>Serum<br>Whole blood |
| MRI | Men with PSA >3 ng/ml and kallikrein score >7.5% | PIRADS score | Digital image |
| Biopsy | Screen-positive men | Post-biopsy symptoms (0, 30 days)<br>Targeted fusion biopsies: number of ROIs, number of biopsies, length of samples,<br>Systemic biopsies: Biopsy length, cancer length and Gleason score per sample, total length of samples, total length of cancer, portion of cancer, global Gleason score, portion of Gleason 4 or 5, perineural invasion | Urine<br>Stool<br>RNA, DNA<br>Cancer tissue and prostate tissue<br>Plasma<br>Serum<br>Whole blood |
| Cancer | Men with | Disease-specific QoL (EPIC-26, MAX-PC) |  |
| **diagnosis** | prostate cancer | Generic Qol/utility (EQ5D, 15D), out-of-pocket costs<br>Gleason/ISUP grade group, number of positive cores, length of cancer, treatment, TNM stage |  |

### Study outcomes and other relevant variables

The primary outcome of the trial is death from prostate cancer. Causes of death will be obtained from the Statistics Finland database and the underlying causes of death will be considered when evaluating if the man died from PCa or from other causes. Cancer cases in the entire trial population including the control arm and non-participants in the screening arm are identified from pathology databases of the two hospitals and through linkages to the Finnish Cancer Registry using the unique PID assigned to all Finnish residents to ensure complete coverage and avoid duplicates (double count).

<u>Secondary outcomes</u> are:

- Diagnosis of prostate cancer (divided into clinically significant and insignificant)
- Progression to advanced prostate cancer (biochemical relapse or progression to metastatic)
- Death due to any cause
- Cost-effectiveness of screening

<u>Adverse outcome variables</u> to monitor screening-related harms are:

- Overdiagnosis of clinically insignificant prostate cancer
- Quality of life impacts of screening and quality of life among men with PCa (EPIC26 instrument)
- Prostate cancer-related anxiety (MAX-PC questionnaire)
- Complications from biopsy (PRECISION questionnaire)

## Statistical analysis

The main analyses will rely on the intention to screen (ITS) principle and will include all randomized men in the two trial arms who were alive and eligible (free of prostate cancer) at the date of randomization. Those men who became ineligible between the date of randomization and first screening invitation will remain in the ITS analysis set.

Two-sided statistical tests will be used, and the overall significance level will be set at 5%. Corresponding p-values will be accompanied with estimates of differences and their 95% confidence intervals.

### Analysis of the primary outcome

The primary outcome of the trial is death from prostate cancer. This is a superiority trial regarding the primary outcome and the comparisons between trial arms will be analyzed and presented on this basis.

Those men who survived will be considered as right-censored observations at the time passed between the time of analysis and time of randomization period. Those men who were lost to follow-up (e.g., due to emigration) will be considered as censored at that particular time (e.g., at emigration). Time to death, defined as the difference between the date of death and date of randomization, will be used as the event time for the analysis.

To evaluate differences between screening and control arms in prostate cancer specific mortality, Gray’s test (Gray 1988) for testing the null hypothesis of equality of cumulative incidence functions will be used. This test differs from the commonly used logrank test in how competing risks of death are treated and is based on the subdistribution hazard of prostate cancer cause of death.

The test will be complemented by reporting the number of PCa deaths, number of men at risk and estimated cumulative incidence functions for each trial arm over follow-up time. The arms will be compared in absolute risks (number needed to invite i.e., the inverse of the risk difference and number needed to diagnose per averted prostate cancer death, i.e., the ratio of excess incidence to mortality reduction), as well as and relative measures of effect (hazard ratios). Descriptive summaries will also be presented by trial centers, age group at randomization.

#### Secondary analyses of the primary outcome

Fine-Gray model for the subdistribution hazard will be used to conduct analyses adjusted for background factors. Outcomes will be compared between age groups and trial centers, and in case of differences, analyses to control for trial center and for age at randomization (categorized as 50–54, 55–59, 60–65 years) will be conducted.

Per protocol analyses excluding men with substantial protocol deviations, such as repeated non-attendance or ineligibility before invitation to screening (with pseudo invitation dates for the control arm), will be conducted if considered pertinent. Additional analyses to correct for contamination and non-compliance, i.e., estimation of efficacy, will be taken by best practices methods at the time of the analyses (e.g., Cuzick et al. 1997).

Descriptive analyses to study effect heterogeneity by center and age group will be performed to complement these analyses. Additional analyses requested by external reviewers or editors in peer-review processes will also be done.

### Analysis of secondary outcomes

#### Diagnosis of prostate cancer

The analysis of cumulative incidence of PCa by disease aggressiveness intends to assess screening impact on detection of clinically significant PCa (representing potential benefit through early treatment) and clinically insignificant PCa (indicating overdiagnosis). The intention is to assess the extent of detection of clinically significant PCa by screening relative to the control arm, and extent of overdiagnosis relative to the control arm. This will inform about the degree of accomplishing rationale of the trial, i.e., detection of aggressive cases at least similar to that in PSA-based screening, while substantially decreasing the yield of low-risk cases. As screening advances the time of diagnosis by several years (lead time), cumulative incidence will be used as the indicator of risk.

Disease aggressiveness will be defined by the International Society for Urological Pathology (ISUP) Gleason grade group. The analyses will be conducted separately for the detection of clinically significant (Gleason 7+ or ISUP 2+) and clinically insignificant (Gleason <7 or ISUP 1) PCa. In secondary analyses, alternative criteria for csPCa will also be employed including ISUP 3+ (Gleason 4+3 or higher), maximum length of cancer tissue in biopsy and number of biopsy cores with cancer.

Risk differences and ratios will be used to infer screening benefits and overdiagnosis compared to the control arm. Besides cumulative incidence, the ratio of aggressive to non-aggressive cases (or proportion of aggressive cancers out of all PCa) will also be reported.

Cumulative incidence for both outcomes will be estimated by trial arm. The overall PCa incidence combines screening benefits and harms and is thus regarded of minor importance in the interpretation of screening impact. Tabular presentations of age at diagnosis, disease stage and grade at diagnosis will be presented.

Both intention to screen (by allocation) and per protocol (screening participants and non-participants) analyses will be conducted for each screening round. For screening participants, screen-detected and interval cases will be reported separately, and screen-detected cases will be broken down by those detected in targeted biopsies of MRI-positive lesions (screening protocol evaluated) and systematic biopsies in screen-negative men with PSA density >0.15 (safety measure to avoid missing clinically significant cases). Any cases detected in a random sample of screen-negative men invited to MRI (analyses to assess underlying prevalence of prostate cancer) will also be reported separately. Analyses to evaluate an optimized screening algorithm will include exclusion of cases with PI-RADS score 3 and kallikrein score calculated also incorporating information on previous biopsies (ignored in the main analysis), as well as using higher cut-off values for PSA and the kallikrein score.

#### Advanced prostate cancer

The analysis of advanced prostate cancer will compare the cumulative incidence of cancer progression, including metastasis and/or biochemical relapse developing after diagnosis and primary treatment, between the screening and control arms. The purpose of the analysis is to evaluate differences between the arms in the risk of developing a potentially lethal, advanced PCa.

The origin of the analysis will be the time of randomization. Cumulative incidence rates will be estimated by the Kaplan-Meier method, and differences between trial arms will be estimated by Cox regression models adjusted by age at diagnosis.

#### Death due to any cause

The analysis of all-cause mortality aims to show that the trials arms are comparable with each other and the general male population in Finland. These analyses will not inform about the effectiveness of screening. Cumulative survival and mortality rates will be estimated by the Kaplan-Meier method, from time of randomization, displayed with frequencies of events and men at risk by trial arm, and by age at randomization.

This analysis will focus on the intention to screen analysis set.

#### Cost-effectiveness

A cost-effectiveness analysis will be performed, incorporating cost data for both out-of-pocket estimated from surveys and service cost data collected from health care providers, as well mortality results (ITS analysis) and utilities based on repeated surveys with 15D and EQ5D instruments (on a random sample of participants). The comparator is no active screening, here represented by the control arm. The main outcome is the incremental cost-effectiveness ratio in terms of costs per quality-adjusted life-year.

A preliminary and exploratory cost effectiveness study can be conducted after the last of the follow-up surveys have been returned, approximately at 3 years after the randomization of the last man into the trial. We plan to undertake a full cost-effectiveness analysis around the time when the evidence on the effectiveness of screening regarding primary outcome has been obtained; this will most likely be near to the analysis at 15 years.

#### Quality of life

These analyses aim to evaluate the short-term and long-term impacts of screening on generic quality of life as well as disease-specific quality of life among men with PCa. Two disease-specific questionnaires, EPIC26 instrument and MAX-PC questionnaires will be used to measure quality of life at 0, 6, 12, and 24 months from PCa diagnosis in both trial arms.

Standard scoring of the EPIC26 instrument will be used. Summary statistics of the five key domains over time and by trial arm will be calculated to assess changes in quality of life of men with PCa from diagnosis onwards. Summary and domain-grouped scores will be analyzed using applications of linear models (or their nonparametric counterparts, if needed) for repeated measures to evaluate differences in quality of life between the arms following PCa diagnosis.

Prostate cancer related anxiety is measured with the Memorial Anxiety Scale for Prostate Cancer (MAX-PC) questionnaire (Roth et al. 2003). Results will be presented as frequencies and percentages for total and subscale scores by trial arm.

Generic quality of life and utilities are evaluated using the 15D and EQ5D instruments as described in the cost-effectiveness section.

### Analysis of adverse outcomes

In addition to detection of low-risk disease by screening as an indicator of overdiagnosis, adverse outcomes mainly relate to the harms due to biopsies. Adverse effects of prostate biopsy are monitored using the questionnaire developed for the PRECISION trial covering pain and other symptoms immediately after biopsy and at 30 days following biopsy. The number of biopsies, as well as the number (%) and type of complications among those with biopsies will be reported.

### Interim analyses and data monitoring

The first analysis of PCa mortality will be conducted at 10 years and the final analysis at 15 years (i.e. at median follow-up time of 10 or 15 years). As we do not intend to stop the trial at 10 years, these interim analyses will be considered as preliminary information. Interim analyses at 10 years will include also analyses of shorter-term benefits.

To control the overall type I error rate (5%) of the trial, we will employ the O’Brien-Fleming rule for alpha spending function. We set the amount of information at 0.5 at 10 years based on the expected numbers of PCa deaths. Thus, by implementation of the O’Brien-Fleming algorithm, the resulting significance level at 10-year interim analysis will be 0.0056, and at the 15-year final analysis 0.0444.

Analyses of secondary endpoints informing about the intermediate outcomes of process indicators including participation, cancer detection, validity and diagnostic performance of the tests in the entire population and subgroups (screened men, non-participants, men in the control arm) will be carried out at regular intervals, as sufficient data become available for evaluation. These will inform about potential need to modify the procedures. Side studies using the samples collected will be carried out to identify new indicators of prostate cancer risk and prognosis.

An <u>independent data monitoring committee</u> (DMC) oversees the trial conduct, and its main task is to ensure safety of the participants. Safety in this context means that screening or screening procedures should not lead to unacceptable disadvantage for the participants in the light of screening benefits. This could take place if the screening intervention had materially worse performance in detecting clinically relevant prostate cancer than anticipated, or substantially higher level of overdiagnosis. The DMC is given a report of the screening results initially every six months and after the first year every 12 months. The DMC can also request any additional information they regard as pertinent to their task. In case of concern, the DMC can recommend discontinuation of the trial; in practice that would mean stopping recruitment and discontinuation of further screening procedures. In addition, they have a mandate to suggest modifications to the trial protocol.

### Handling of missing data

Extent of missing data will be described, for example, by presenting the number of individuals with missing values per variable.

For outcome variables relying on dates – dates of randomization, censoring, diagnosis or death – incomplete dates will be imputed by 15 (in the case that the day variable was missing, but known month and year), and by 30/6 (in the case that only the year was known).

In case a substantial proportion of men (at least 5%) have missing data on one or more variable needed for the effectiveness analysis in question, multiple imputation methods will be used to demonstrate the robustness of findings (Little *et al*. 2012). Imputation models will include outcome variables and trial arm in addition to all variables relevant to the particular analysis. Final estimates will be derived by combining estimates and their standard errors across data sets using Rubin’s rules.

### Data management and quality assurance

RedCap database application is used for data management in the trial, covering all major data types from questionnaires and lab results to MRI findings, diagnoses and causes of death. RedCap allows access defined by two-factor authentication (2FA) and flexible definition of user-specific functions and rights.

In REDCap, variable specific parameters and predetermined options are used to prevent entering invalid data (e.g. predefined values and acceptable ranges). All data is verified from the original data source and monitored monthly. Until the verification, data is saved as incomplete or unverified. Lead times between screening tests are monitored every 6-8 weeks. For the laboratory work (including sampling, processing, and storing) each task has a protocol shared by the study centers. Any deviations from the sample specific protocol are documented.

## Conclusion

This statistical analysis plan lays out the plans for outcomes of the trial, including the definitions of important outcomes, analysis principles and interpretation, methods for primary analysis, pre-specified subgroup analysis, and secondary analysis.

## Data Availability

All data produced in the present work are contained in the manuscript

## Authors’ contributions

All authors approved the final version of this manuscript. Study concept and design: AA, AR, JN, JR, KN, TK, TT; drafting of the manuscript: AA, JN; critical revision of the manuscript for important intellectual content: AA, AR, JN, JR, KN, TK, TT.

